# Asymptomatic *Bordetella pertussis* infections in young African infants and their mothers identified within a longitudinal cohort

**DOI:** 10.1101/2020.11.18.20231423

**Authors:** CJ Gill, CE Gunning, W MacLeod, L Mwananyanda, D Thea, R Pieciak, G Kwenda, Z Mupila, P Rohani

## Abstract

Despite long-standing vaccination programs, pertussis incidence has increased in numerous countries; transmission by asymptomatic individuals is a suspected driver of this resurgence. However, unequivocal evidence documenting asymptomatic infections in adults and children is lacking due, in part, to the cross-sectional nature of most pertussis surveillance studies. In addition, modern pertussis surveillance relies on quantitative PCR (qPCR) using fixed diagnostic thresholds to identify cases.

To address this gap, we present a longitudinal analysis of 17,442 nasopharyngeal samples collected from a cohort of 1,320 Zambian mother/infant pairs. Using full-range cycle threshold (CT) values from IS481 qPCR assays, we document widespread asymptomatic infections among mothers and also, surprisingly, among young infants. From an initial group of eight symptomatic infants who tested positive by qPCR, we identify frequent contemporaneous subclinical infections in mothers. Within the full cohort, we observe strong temporal correlation between low- and high-intensity qPCR signals. We compute a single time-averaged score for each individual summarizing the evidence for pertussis infection (EFI), and show that EFI strongly clusters within mother/infant pairs, and is strongly associated with clinical symptomatology and antibiotic use.

Overall, the burden of pertussis here is substantially underestimated when restricting diagnostic criteria to IS481 CT≤35. Rather, we find that full-range CT values provide valuable insights into pertussis epidemiology in this population, and illuminate the infection arc within individuals. These findings have significant implications for quantifying asymptomatic pertussis prevalence and its contribution to overall transmission. Our results also expose limitations of threshold-based interpretations of qPCR assays in infectious disease surveillance.

**IMPORTANCE STATEMENT:** Current pertussis epidemiology rests largely on cross-sectional surveys that use diagnostic thresholds to interpret qPCR results as positive or negative, and thus fail to capture arcs of infection within individuals or populations. By longitudinally monitoring a cohort of African mother/infant pairs and using full-range qPCR results, we quantify the otherwise-hidden evidence for pertussis infection (EFI) in individuals. We demonstrate strong clustering of EFI within mother/infant pairs and quantify the association between EFI and both pertussis symptoms and antibiotic use. Critically, we find strong evidence that asymptomatic pertussis is common in both infants and mothers, indicating that the burden of pertussis has been significantly underestimated in this population. Our results also inform qPCR-based monitoring of other pathogens, such as SARS-CoV-2.

## INTRODUCTION

*Bordetella pertussis* remains a significant cause of morbidity and mortality among infants and young children around the world,^1,2^ and has experienced a resurgence in numerous countries despite long-standing vaccination programs^3-6^. Transmission by asymptomatic individuals is a suspected driver of pertussis resurgence,^7,8^ though unequivocal evidence documenting asymptomatic infections in adults and children is lacking. Rapid and reliable molecular diagnosis of pertussis is now possible using quantitative PCR (qPCR), which has supplanted microbiologic culture as the preferred tool for detecting pertussis.^9^ Yet most pertussis surveillance uses cross-sectional monitoring that only captures a single point in time and thus cannot distinguish asymptomatic from pre-symptomatic infections.

In principle, repeated sampling over time could unambiguously identify asymptomatic infections. For example, if one could identify a patient at the moment of pertussis exposure and then regularly monitor them over time, one could anticipate capturing a gradual ‘fade-in/fade-out’ sequence as bacterial load, and thus qPCR signal intensity, varies across the arc of the infection: at initial exposure (where bacterial density is below the assay’s limits of detection), during acute infection (as bacterial density—and signal intensity—rises to a peak), and finally during recovery and convalescence (where an eventual loss of signal indicates pathogen clearance). Indeed, recent human infection trials have borne out this scenario.^10,11^ By sampling a single point in time, however, cross-sectional observations lack the historical context necessary to trace this arc of infection.

An additional complication arises when diagnostic thresholds are applied to qPCR cycle threshold (CT) values to distinguish positive and negative cases. For example, an IS481 CT<35 has been used as a diagnostic threshold of pertussis.^12,13^ This process introduces several problems, including calibration (e.g., between labs, machines, or over time) and a poorly examined trade-off between sensitivity and specificity. Further, this process discards information about qPCR signal intensity, such that borderline or low-intensity signals are summarily discounted as false positives or ‘indeterminate’. These complications become particularly important as we extend the use of qPCR from clinical diagnosis into disease surveillance of populations.^14^

Here we present an analysis of 17,442 nasopharyngeal samples (and associated IS481 qPCR assays) collected from 1,320 Zambian mother/infant pairs who each provided at least 4 samples during the study. We begin with a descriptive analysis of eight mother/infant pairs where each symptomatic infant had definitive qPCR-based evidence of pertussis infection. We document the arc of infection in these individuals and observe frequent contemporaneous subclinical infections in the mothers of these infected infants. We then turn our attention to the entire cohort, where we use full-range IS481 CT values to show that qPCR signals of different intensities cluster in time, and quantify the evidence for pertussis infection (EFI) within individuals. We show that EFI strongly clusters within mother/infant pairs, and is strongly associated with clinical symptomatology and antibiotic use.

In total, we find that full-range CT values yield valuable insights into pertussis epidemiology in this population. Critically, the burden of pertussis here is substantially underestimated when restricting diagnostic criteria to IS481 CT≤35. We also demonstrate widespread asymptomatic pertussis infections among mothers and, surprisingly, among young infants.

## RESULTS

### Study overview

In 2015, we partnered with the Bill & Melinda Gates Foundation to conduct a prospective cohort study in Lusaka, Zambia.^15^ Between April and November 2015, we enrolled 1,981 healthy Zambian mother/infant pairs (3,962 individuals) shortly after birth and observed them during an additional six clinic visits scheduled at roughly 2-3-week intervals through approximately 14 weeks of age (when the last of the three routine infant DTP vaccine visits occurs). At each visit, we systematically obtained nasopharyngeal (NP) swabs and assessed symptoms and antibiotic use.

Of the initial cohort of 1,981 pairs, 1,497 mother/infant pairs attended at least one post-enrollment clinic visit, and 834 pairs attended all seven scheduled visits (including enrollment, **Figure S1-S2**). In this analysis, we focus on the 1,320 pairs with ≥4 NP samples per subject (**Figure S3**). Baseline cohort demographics are shown in **Table 1**. Infants were enrolled at a median of 7 days post-partum; 53% were male, with a median gestational age of 40 weeks and birth weight of 3000 grams. Mothers’ median age was 25 years; >90% were married, and 17.5% were known to be infected with HIV. Among the HIV positive mothers, nearly all were on antiretroviral therapy (ART) at the time of enrollment, and half had initiated ART prior to conception. Nearly all mothers received at least one dose of tetanus toxoid during pregnancy, signaling that some antenatal care was received by at least 99.5% of the maternal cohort.

**Table 1:** Demographic characteristics of participants (interquartile range in parentheses). Only subjects with at least 4 NP samples were included in subsequent analyses.

| Parameter | Study Participation |  |
| --- | --- | --- |
| | Enrolled | $\geq 4$ NP Samples |
| Number Under Study | 1981 | 1320 |
| **Mothers** |  |  |
| Married | 90.2% | 89.8% |
| HIV+ | 17.5% | 19.5% |
| Median Age | 25 (21, 29) | 25 (22, 30) |
| Median Infants In House (<1 year) | 1 (1, 1) | 1 (1, 1) |
| Median Children In House (< 5 years) | 2 (1, 2) | 2 (1, 2) |
| **Infants** |  |  |
| Born at Chawama PHC | 56.9% | 56.6% |
| Born at UTH | 34.8% | 35.5% |
| Female Sex | 46.9% | 46.1% |
| Median Birth Weight (kg) | 3 (2.8, 3.3) | 3 (2.8, 3.3) |

### Descriptive analysis of eight noteworthy mother/infant pairs

**Figure 1** provides a detailed timeline of IS481 CT results from the initial group of eight mother/infant pairs, which included all infants with a definitive positive NP sample (IS481 CT<35) during a symptomatic visit. This figure highlights the experimental design, where pairs were monitored across the infant’s first months of life at regularly spaced clinic visits (e.g., pairs C & E), with additional mother-initiated visits for acute health care (e.g., pairs A & D). Minimal symptoms (cough and/or coryza) were common in both mothers and infants, while moderate to severe symptoms were much rarer, particularly among the mothers. Also rare is ptxS1 corroboration of IS481 results (pair D and infants F-H). This is not surprising, given the high IS481 copy number relative to the single ptxS1 copy, which renders the latter target specific but insensitive for detection of the pathogen.^16^

**Figure 1:**
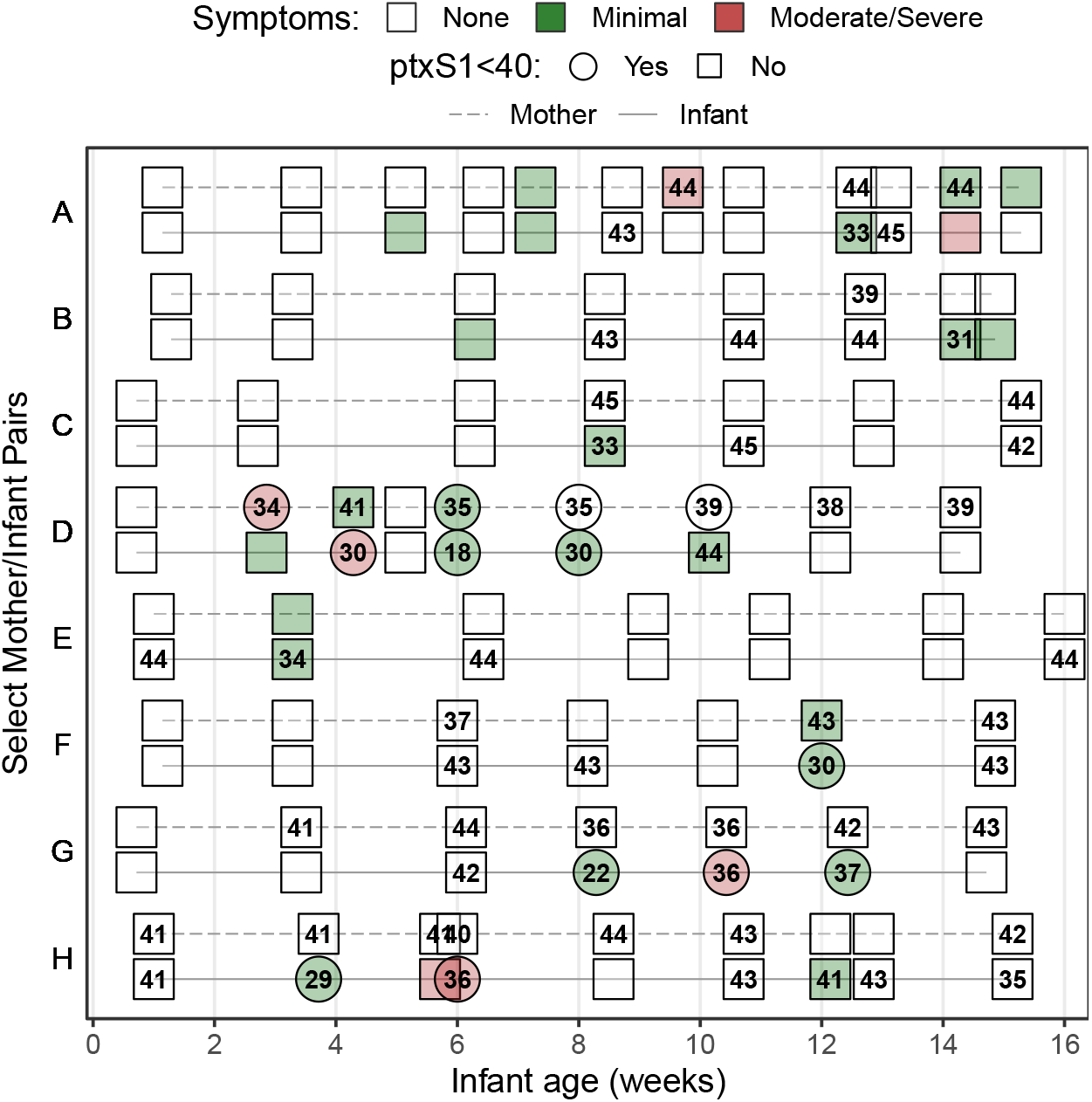
Select mother/infant pairs over time, showing rounded IS481 Ct values (numbers), ptxS1 results (shape), and pertussis symptoms (color) at each clinic visit; selection includes all symptomatic infants with definitive evidence of pertussis infection (IS481<35). Blank cells show NP samples with no detected IS481. Contemporaneous detection of IS481 within pairs is common, as are temporal clusters of IS481 within individuals. Pertussis symptoms are relatively uncommon in mothers; indeed, 4 out of 7 mothers with detectable IS481 presented no pertussis symptoms.

Of particular note, seven of these eight mothers had at least one detecting assay, while six had multiple detections. However, many of these mothers’ detecting assays showed relatively weak signals (IS481 CT>40, e.g., mothers A, C, and H), with only a few results meeting CDC’s recommended diagnostic criteria for a positive test.^17^ We note, however, that CDC criteria were designed for application by clinicians when evaluating patients with severe and/or classic pertussis symptoms, and were intended to favor specificity over sensitivity. They were not designed for surveillance of asymptomatic individuals. Moreover, these weak qPCR signals frequently bracketed visits with definitive test results and were observed in all eight infants who met CDC criteria at one or more visit.

A clear example of pertussis infection fade-in and fade-out is provided by infant G, including ptxS1 confirmation and severe symptoms that occur after the observed IS481 signal peak. Remarkably, a weak (and asymptomatic) IS481 signal in infant G at age six weeks precedes the acute infection observed around age 8 weeks, while a weak IS481 signal in mother G is observed even earlier, before 4 weeks of age. This example suggests the likely sequence of transmission events within this mother/infant pair and thus influenced our emerging conclusion that weaker IS481 signals should not be automatically discounted.

Mother H provides another illustrative example, where seven of nine assays (78%) detect IS481, but none reach the canonical diagnostic threshold of <35, and none had a detectable ptxS1 result. *A priori*, the probability that all of these detections were false positives appears low. In the context of an infected infant, this explanation becomes even less likely. While low sample pathogen density could also reflect clinically uninformative variation in sample collection and processing, the more parsimonious interpretation is that these seven NP samples contained pertussis, albeit at a low density.

While all infants in this first analysis were selected based on their presentation of symptomatic pertussis, several of their mothers presented with no respiratory symptoms at clinic visits. Of particular note are mothers G and H, each of whom had multiple detecting assays, strongly suggesting asymptomatic (or minimally symptomatic) pertussis infections.

To assess how frequent low-intensity signals were in our cohort, 500 NP samples were randomly selected and tested from our catalog of over 9,000 maternal samples, none of which yielded detecting results. Applying the binomial theorem for an expected frequency of just under 1/500 (i.e., assuming that the next sample would have been detecting), the probability that seven of eight mothers of infected infants also had one or more detecting assays occurring by chance was <0.0001. We conclude that random chance is unlikely to account for the high concordance within these pairs, or for their evident tendency to coincide in time.

### Quantitative analysis of the full cohort

The low-intensity IS481 CT values (i.e., ≥35) discussed above would be adjudicated as ‘negative’ or ‘indeterminate’ in a typical cross-sectional study. However, we observed multiple lines of evidence supporting their microbiological and epidemiological significance that compelled us toward a comprehensive analysis of full-range IS481 CT values for the entire cohort.

#### Concordance of qPCR signals over time

If weak qPCR signals (e.g., CT≥40) represent random background noise (i.e., false positives), then we would anticipate random variation in their frequency over time, and no evidence of correlation with stronger signals. To test this hypothesis, we assess the relative frequency of four strata of qPCR signal intensities over time. In **Figure 2A**, the time-averaged frequency of each stratum is shown by a separate curve, where each study participant could contribute multiple detecting assays to each curve. We also show individual high-intensity assays (CT<35) as dots. Here we observed a strong correlation among all strata over time in both mothers and infants, with a peak in June and July and a long tail of decline in September and October.

**Figure 2:**
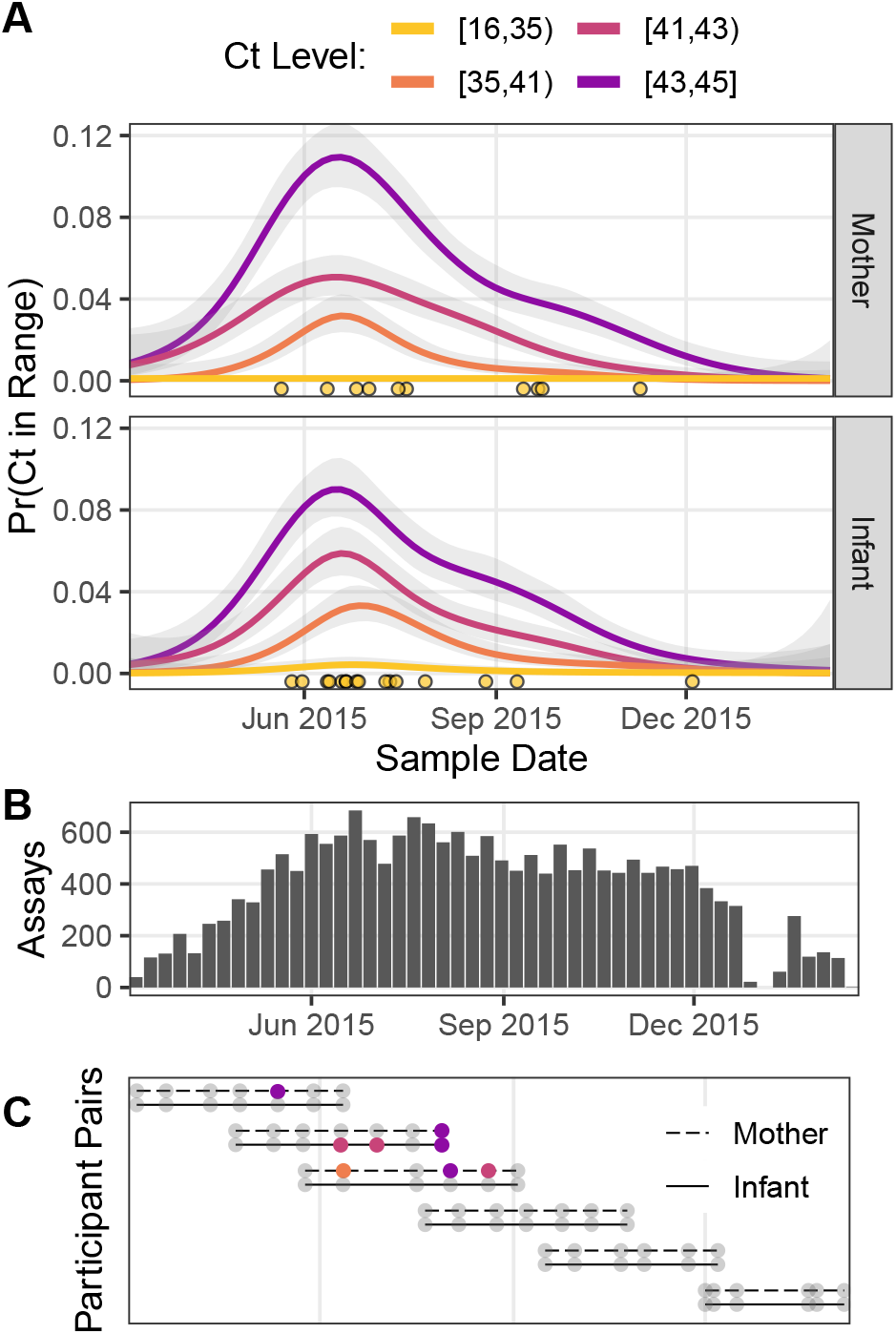
IS481 qPCR assays over time. **A**, proportion of detecting assays over time stratified by signal intensity (estimated via GAM smoother), where lower Ct values indicate more IS481. Points show assays with Ct<35. A cluster of detecting assays in all strata peaks in late June / early July. The temporal correlation observed among strata is consistent with detection of a pertussis outbreak, but is not consistent with randomly distributed false positive assays. **B** shows the number of assays per week (denominator of A). The dip in Jan 2016 corresponds with the Christmas holiday. **C** illustrates study participation for a representative sample of mother/infant pairs, showing clinic visits over time (color as in A). Visits included initial enrollment (shortly after birth) followed by (up to) 6 scheduled visits at 2-3 week intervals, and (in some cases) additional mother-initiated visits.

In our published 2016 analysis, we had identified a cluster of definitive infant pertussis cases between May and July of 2015 that contained the eight infants described above.^15^ While assays with CT<35 were few (yellow points in **Figure 2A**), even the weakest intensity stratum (e.g., CT>43), which was also the most frequent, closely mirrored and possibly preceded in time the higher intensity strata (e.g., CT<41). We also note that cohort size alone cannot explain these results, as the cohort’s size reached a steady state in June of 2015 that was sustained through the end of December 2015 (**Figure 2B**). Rather, these results are consistent with a population-level ‘fade-in/fade-out’ dynamic, where multiple overlapping signals from single individuals (**Figure 2C**) create these curves over time. This also highlights the preponderance of detecting assays with low signal intensity. We note that the range of CT values appeared greater for infants than mothers, with twice as many with CT results below 35 (15 vs 8, RR 0.54 95% CI 0.2-1.3).

Plotting these results by infant age also showed a gradual increase in detecting assays over time in infants but not mothers (**Figure S4**). The breakdown of IS481 results is summarized in **Table S1**. Approximately 91% of all tests were negative (non-detecting), whereas only 0.11% and 0.18% of mother and infant samples, respectively, had CT<35 and would have been considered definitive positive samples; all other samples would have been deemed indeterminate or negative based on traditional cut-points.

#### Quantifying evidence for pertussis infection

Our next analysis combines the contextual information provided by repeated sampling with that of full-range IS481 CT values to quantify the evidence for pertussis infection within individuals. Here we focus on the 1,320 mother/infant pairs where 4 or more NP samples (and associated IS481 assays) were available for each subject (see also **Figure S3**). For illustrative purposes, we also provide individual results from the following analyses for the eight mother/infant pairs described above in **Table S2**.

We first compute the reverse cumulative distribution (RCD) plots for CT values for mothers and infants separately (**Figure 3A**). We then use these RCD plots to compute, for each subject, a summary statistic that we term the “evidence for infection” (EFI): one minus the geometric mean RCD probability. Here, EFI=0 indicates no evidence (no detecting assays), while an EFI approaching 1 indicates very strong evidence arising from more detecting assays and/or stronger signals. We note that by averaging across time EFI provides no information about when infection occurred within the study. In **Figure 3B**, we show the distribution of EFI in mothers and infants stratified by the number of detecting assays. Under the premise that many false positives would be highly unlikely regardless of signal intensity, we categorize individuals with three or more detecting assays as having *strong EFI*, as well as other individual within this EFI range (dotted line, EFI>=0.52). Individuals with intermediate evidence (0<EFI<0.52) are categorized as having weak EFI.

**Figure 3:**
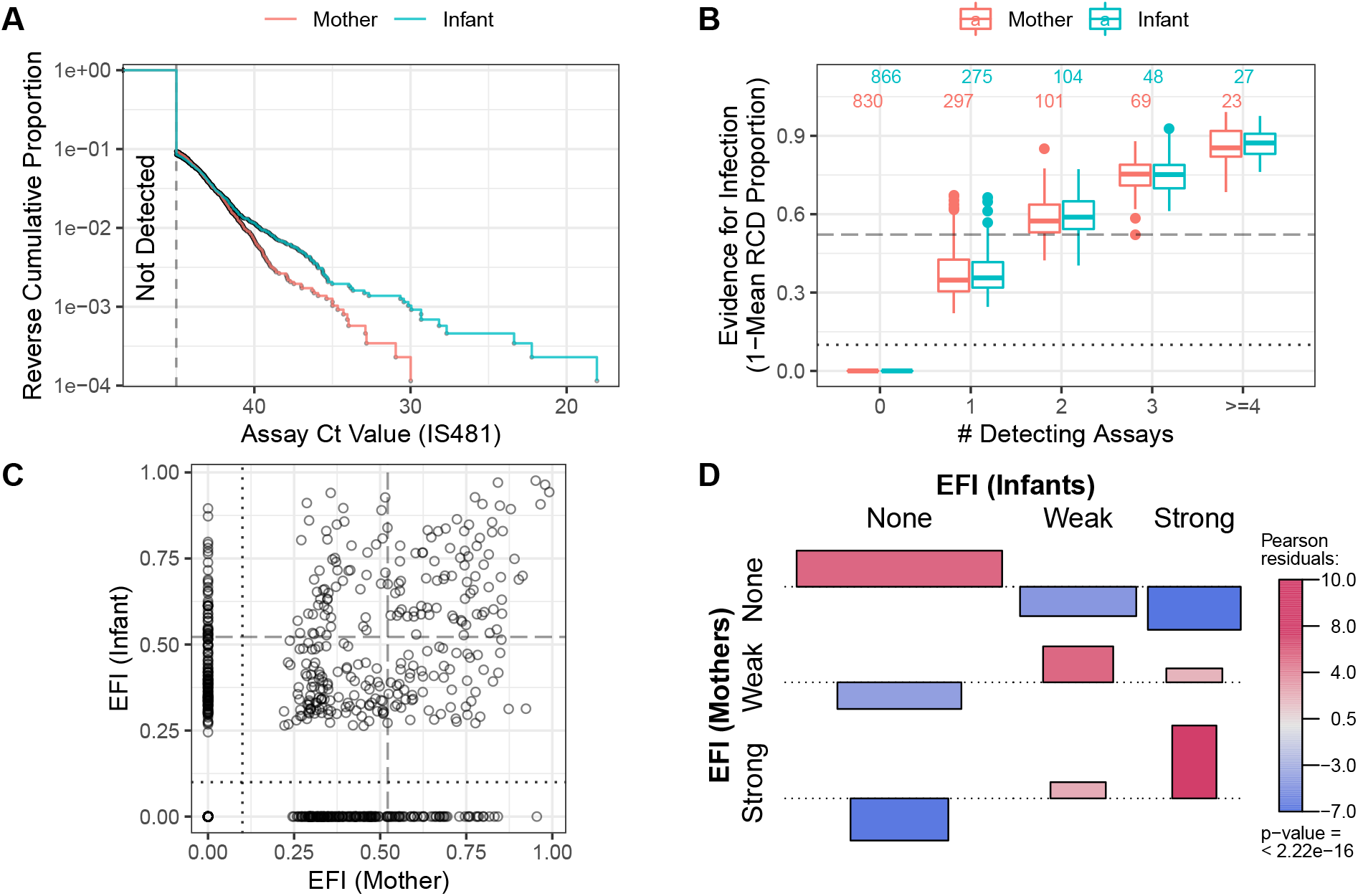
Quantifying evidence for pertussis infection, and concordance of evidence within mother/infant pairs. **A**. Reverse cumulative distribution (RCD) curves of IS481 Ct values for mothers and infants. **B**. Boxplot summarizing evidence for infection (EFI), stratified by number of detecting assays per subject (x-axis). For each subject, EFI equals one minus the geometric mean RCD proportions (as in A). In general, EFI increases with lower Ct values (A) and more detecting assays. The dashed line delineates strong evidence (defined to include all subjects with ≥ 3 detecting assays, 0.52 ≤ EFI<1) from weak evidence (0<EFI<0.52); dotted line delineates no evidence (EFI=0). **C**. EFI in mother/infant pairs. Dotted and dashed lines as in B for mothers (vertical) and infants (horizontal). **D**. Association of EFI strength (from C) between mothers and infants, showing very strong concordance (red) and rare discordance (blue) within pairs, particularly for pairs exhibiting strong EFI. Bar widths are proportional to expected counts; bar height and color show Pearson residuals (scaled difference between observed and expected counts). P-value and residuals are relative to independent association. See also Figure S5.

#### Concordance of evidence within mother/infant pairs

Multiple prior studies have reported that mothers and close family contacts of infected infants are very likely to also be infected,^18-20^ as consistent with our results in **Figure 1**. Here we use EFI to assess concordance of infection status within mother/infant pairs across the study. In **Figure 2C**, we show the intersection of EFI scores in each mother/infant pair as a single point. We use dotted and dashed lines (EFI>0 and EFI>0.52, respectively) to delineate possible EFI combinations for each mother/infant pair, e.g., mother-weak/infant-none or mother-strong/infant-strong. **Figure 2D** highlights the strong association of EFI category within mother/infant pairs. This association plot shows the frequency of concordance relative to expectation (assuming independent assortment), plotted as Pearson residuals. We find that when infants have no EFI, the corresponding mothers’ EFI is also likely to be absent. Conversely, when infants display strong EFI, evidence in mothers also tends to be strong. We also repeat this analysis by varying the threshold defining a strong EFI score (using either two and four detecting assays, respectively), and find consistent results throughout (**Figure S5**). While this analysis does not provide evidence of *contemporaneous* infection within pairs, these results offer strong evidence of transmission within the mother/infant pair.

#### Concordance of evidence with symptoms

Assuming that greater EFI represents a higher bacterial burden over time and thus yields a higher likelihood of symptomatic illness, we tested whether EFI category was associated with cough and/or coryza (minimal symptoms), or additional pertussis symptoms (moderate to severe symptoms). In **Table 2**, we tabulate the frequency of EFI category stratified by symptoms for mothers and infants. As shown in **Figure 4A**, among infants there is a strong positive relationship between EFI and the presence and severity of respiratory symptoms. A similar pattern is observed in mothers, except that moderate to severe symptoms were rare in mothers (**Figure 4B**). This absence of symptoms in mothers across infection status (i.e., for both weak and strong EFI) is consistent with the well-documented protective effects of pre-existing immunity.^21^ On the other hand, we observe 41 infants with strong EFI who were asymptomatic at all study visits (**Table 2**). While these infants are much less common than expected by random chance (**Figure 4A**), they nonetheless represent more than 3% of cohort infants.

**Table 2:**
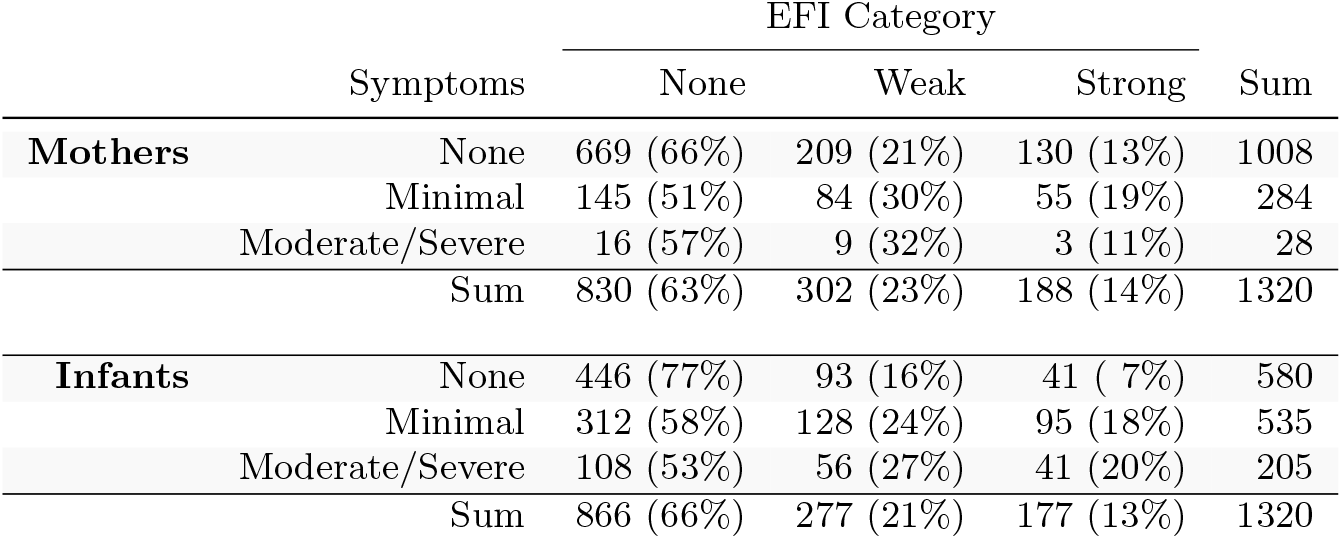
Frequency of EFI category in mother and infants, stratified by occurrence of symptoms at any point during study participation. Percentages are relative to row sums. Minimal symptoms include coryza and/or uncomplicated cough. Moderate to severe symptoms include all other pertussis symptoms in the Modified Preziosi Scale.

**Figure 4:**
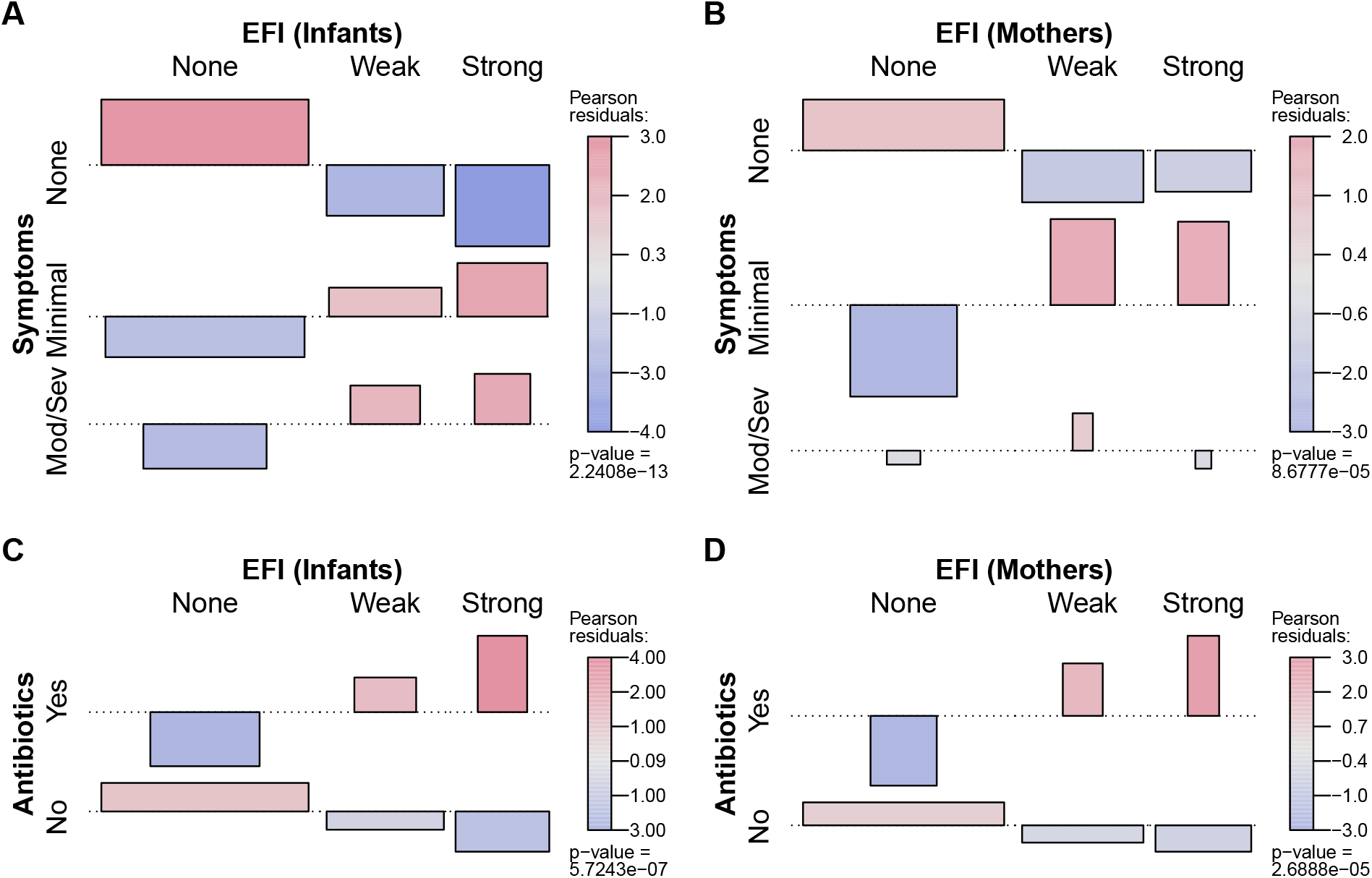
Association between participant EFI (as in Figure 3) and documented pertussis symptoms (**A-B**) or antibiotic use (**C-D**), separating mothers and infants (columns). Minimal symptoms include cough and/or coryza only; bar heights and p-values as in Figure 3D. In A-B, frequent co-occurrence of strong EFI with minimal symptoms is evident in both mothers and infants, as is no symptoms with no EFI. In infants, moderate to severe symptoms commonly co-occur with strong EFI, while more severe symptoms are rare in mothers. In C-D, frequent co-occurrence of antibiotic use with strong EFI is evident in both mothers and infants.

#### Concordance of evidence with antibiotic use

Finally, we observe a strong positive association between EFI and antibiotic use in both infants and mothers (**Fig 4C-D**). We cannot infer a causal direction from the observed association. On one hand, antibiotic use presumably serves as a proxy for symptoms at some prior time, since a clinician must have judged the individual sick enough to warrant treatment. On the other hand, antibiotic use has been shown to reduce the infectious dose of *B. pertussis* through effects on respiratory microbiota^22^. We also note that amoxicillin, which is not effective against *B. pertussis* but is a first-line therapy for empiric treatment of pneumonia, accounted for the vast majority of antibiotics use in this study. Erythromycin, which would be active against pertussis, is available in Zambia but is not part of the Integrated Management of Childhood Illness algorithm by which children are diagnosed with pneumonia, and hence rarely used. With the above in mind, our results call into question whether ineffective antibiotics have been commonly prescribed for mild pertussis cases in this (and possibly other) populations.

## DISCUSSION

Our initial investigation of eight noteworthy mother/infant pairs suggested that full-range IS481 CT values contained useful information about the pertussis infection process. Building on this, we conducted a comprehensive analysis of our extensive library of NP samples and directly-observed clinical records that were prospectively collected from a representative African urban cohort of Zambian mother and infants. We find that the true burden of pertussis infection in this population was much higher than we expected, most of which reflected asymptomatic or minimally symptomatic infections. We also observe clustering of qPCR signals over time consistent with a local outbreak, and clustering of detecting assays within mother/infant pairs. Finally, we observe a strong association between the pertussis EFI and pertussis symptoms and the use of antibiotics. Collectively these findings indicate that our longitudinal analysis of unthresholded qPCR data provided valuable and novel insights into the scope and scale of pertussis transmission in this population.

A key finding of our investigation is the common occurrence of asymptomatic (and/or minimally symptomatic) pertussis infections in adults. This issue has long been a subject of debate and speculation but has remained unresolved due to the lack of longitudinal data sets that would allow one to distinguish asymptomatic from pre-symptomatic infections.^7,23-27^ Far more surprising, however, is the relatively high frequency of asymptomatic infections among infants. Pertussis in infants has been assumed to be predominantly severe and occasionally mild; our data indicate otherwise.^28-30^

Older children, adolescents and young adults are generally recognized as the reservoir for pertussis,^31-34^ while mothers and older siblings are often the source of pertussis in infants.^35-38^ While the relationship between qPCR signal intensity and infectiousness may be modified by other factors, including whether the individual is symptomatic or whether *B. pertussis* is growing freely or embedded within a biofilm matrix,^39^ signal intensity nonetheless serves as a rough proxy for infectiousness.^40-43^ In our study, the distribution of IS481 CT values were similar for infants and mothers, raising the question of whether infants were merely the recipients of pertussis infections, or whether they could also participate in chains of transmission.

This analysis has several limitations. We have not provided a formal analysis of the related species *B. parapertussis* or *B. holmesii*, though initial estimates suggest that neither plays a major role in this system. We have not directly inferred pertussis transmission, and acknowledge that a linear relationship may not exist between qPCR signal intensity and infectiousness. In addition, several key questions fell outside of the scope of this analysis and warrant further exploration. Notably, a time-series analysis of the trajectory of qPCR signals within individuals over time, and within mother/infant pairs, remains an important next step, as does a formal calculation of the incidence of symptomatic and asymptomatic pertussis. Finally, an analysis of the relationship between qPCR signal intensity during infection and the administration of routine DTP vaccinations to study infants is also warranted.

A key strength of our study is that our data were systematically collected according to a schedule rather than adventitiously sampled in response to symptoms. Such prospective surveillance ensures these data are robust against selection bias, which is the primary threat to any cohort. Moreover, we had a high retention rate (Gunning et al, BMJ Open, In Review), and the majority of retained participants attended all seven scheduled visits. During the study, our team offered convenient access to participants’ acute care needs, as well as travel stipends at each visit, thus reducing the likelihood that infection events were missed due to patient seeking care elsewhere. In addition, automated sample tracking allowed us to link qPCR results directly to concurrently measured respiratory symptoms and clinical data including antibiotic use, which greatly increased our confidence that we could sort symptomatic from asymptomatic infections.

We note that *B. pertussis* has proved to be a valuable study organism for several reasons. First, IS481 provides a very sensitive target due to its high copy number, while *B. pertussis* can be easily collected from the nasopharynx. In addition, pertussis infections can last for many weeks (or even months), permitting a sampling frequency on the order of once every few weeks. By contrast, the arc of most viral respiratory infections is measured in days (or a few weeks), and more frequent sampling would be required to observe these shorter arcs of infection. Nonetheless, the general principles outlined here remain salient: that a weak signal tracked over time can nonetheless provide valuable insight into biological processes. Even in situations where repeated, longitudinal sampling of individuals is not feasible, a careful analysis of full-range qPCR data could be applied to cross-sectional samples that were prospectively collected from a population over time. In the same way that overlapping qPCR signals across the full range of intensity appear to expand the apparent duration of the pertussis outbreak detected in our study, an important outstanding question is whether a sustained population-level rise in low-intensity qPCR signals could provide early warning for epidemics of RSV, influenza, or CV19. Indeed, there is an emerging dialog about the use of full range qPCR data as tools in epidemiologic surveillance.^44,45^

In conclusion, these results provide an important benchmark of the relative frequency of asymptomatic and minimally symptomatic pertussis infections in both adults and infants. More broadly, the longitudinal structure of our data suggests that the use of canonical diagnostic thresholds in qPCR (e.g.IS481 CT<35) has the unintended effect of removing valuable epidemiological information, which is particularly impactful of population-level surveillance. Our findings also have implications across a broad range of infectious disease surveillance efforts, where we believe a similar approach could potentially provide early warning of infectious disease outbreaks. Future analyses in our group will focus on quantifying the overall burden and the relative contribution to transmission of symptomatic and asymptomatic pertussis in infants and mothers separately, exploring the direction of transmission within mother/infant pairs, and using these data to update models of pertussis transmission.

## MATERIALS AND METHODS

### Study Design

The Southern Africa Mother Infant Pertussis Study (SAMIPS) was a longitudinal cohort study conducted in Lusaka, Zambia that followed mother/infant pairs through the infants’ first three months of life. To this end, we sought to enroll all healthy live births that occurred between March and December 2015 in Chawama compound, a densely populated peri-urban slum near central Lusaka. A detailed account of study methods (including sample size considerations) was published previously.^15^

Enrollment was conducted at the Chawama Primary Health Clinic (PHC), and mother/infant pairs were recruited during their first scheduled postpartum well-child visit at approximately 1 week of age. Chawama PHC is the only government-supported clinic in this community, and is the primary source of medical care for Chawama residents, allowing us to maximize study reach. Prior to study initiation, a public outreach campaign also provided study information to pregnant Chawama residents.

Infant enrollment eligibility included the following: full-term (infants were born after 37 weeks), birth weight >2500 grams, and delivered without complications or apparent disease. Maternal eligibility included signed consent, Chawama residency (anticipated remaining in the community during study period), known HIV status prior to delivery, and treatment with prophylactic antiretroviral therapy at the time of delivery for HIV+ mothers.

Mothers were incentivized to join and remain in the cohort in three ways. First, the SAMIPS medical staff provided all routine and acute medical care for study participants during their time of enrollment. This significantly reduced waiting times for care from over 3 hours to half an hour or less. Second, mothers received a travel stipend for each visit valued at approximately 7 US dollars. Lastly, a small gift of baby supplies was provided for mothers attending the final scheduled study visit.

After the baseline enrollment visit, infants were scheduled for six routine follow-up clinic visits at 2-3-week intervals through approximately 14 weeks old (maximum, 18 weeks). Additional unscheduled clinic visits were initiated by study mothers for acute medical care as well as routine well-child care. At each clinic visit, nasopharyngeal (NP) swab samples were obtained from both mother and infant, and detailed records of current respiratory symptoms were collected on a standardized reporting sheet for each subject by clinic staff. Unique barcodes were assigned to study records from pre-printed sticker books, and were used to link subjects, clinic visit records, and NP samples. Each barcode was scanned at the time of assignment using the Xcallibre digital pen system. We note that qPCR results were unavailable during the study, and thus could not affect symptom assessments and clinical management decisions.

Routine childhood vaccinations were administered during scheduled clinic visits. Diphtheria-Tetanus-Pertussis (DTP) doses 1-3 was administered at visits corresponding to 6, 10, and 14 weeks of age as a pentavalent combination (Pentavac, Serum Institute of India Limited, Pune, India) that included whole-cell pertussis, *Haemophilus influenzae* type B (HIB), and Hepatitis B. The pneumococcal conjugate vaccine (PCV10) was co-administered with DTP, and the oral rotavirus vaccine administration was scheduled for 6 and 10 weeks of age. Additional details regarding infant vaccination are provided in Gunning et al (BMJ OPEN 2020, under review). Routine childhood vaccinations were provided by regular clinic staff at no cost in a separate area of the clinic compound located ∼50 feet from where the NP sampling was done. This was intended to avoid contamination of the swabs by pertussis DNA present within the pertussis vaccines themselves, a known cause of pseudo-outbreaks of pertussis in health care settings.^46^

### Nasopharyngeal sampling

NP samples were obtained using flocked-tipped nylon swabs (Copan Diagnostics, Merrieta, California)^47^ that were inserted into each nostril until contact with the posterior nasopharynx was made. Swabs were then rotated 180 degrees in both directions, placed in commercially prepared tubes with universal transport media (UTM), and stored on ice until transport. Samples were collected from the study clinic daily and were taken to the PCR laboratory at the University Teaching Hospital (UTH), where they were stored at −80°C.

### Laboratory methods

NP sample DNA was extracted using the NucliSENS EasyMag system (bioMérieux, Marcy l’Etoile, France).^48,49^ Samples were initially tested for *B. pertussis* using a singleplex TaqMan qPCR genomic assay targeting the IS481 insertion sequence. In addition, a qPCR assay tested each sample for the constitutively expressed human RNase P (RNP) to assess successful sample collection, storage, DNA extraction, and lack of PCR inhibition. Each 96-well qPCR plate contained approximately 46 samples (one each of IS481 and RNP), along with a positive and negative control for each assay. We note that a lower CT value indicates a greater quantity of target. Each reaction was run for 45 cycles, such that the minimum detectable target quantity has a CT value of 45. We consider assays with a CT value of 45 or less to be *detecting* assays; all others are *non-detecting* (N.D.) assays.

For the descriptive analysis of the first eight symptomatic infants and their mothers, an IS481 detection was followed by a second assay targeting the pertussis toxin gene ptxS1. Given the high volume of testing, we only used IS481 for the full analysis of the library. All primers and probes were purchased from Life Sciences Solutions (a subsidiary of ThermoFisher Scientific Inc). Most samples were run using an ABI 7500 thermocycler (ThermoFisher Scientific Inc, Waltham, MA). Starting in 2019 some samples were also run on a QuantStudio5 thermocycler (ThermoFisher Scientific Inc, Waltham, MA). An analysis of samples run in parallel on both machines showed minimal systematic variation between machines, so we do not distinguish these in the current analysis.

### Data and Statistical Analysis

#### Descriptive analysis of the first eight mother/infant pairs

As presented in our 2016 paper, from the 1,981 infants in SAMIPS, we initially selected infants presenting with any respiratory symptoms (rather than classic pertussis symptoms) for PCR testing.^15^ Thus, any pertussis detected in these infants will be ‘symptomatic pertussis’ by definition. Given our focus at this stage on detecting symptomatic pertussis, we defined pertussis strictly as per the US CDC’s protocol: any IS481 insertion sequence CT <35, or an IS481 of 35-40 plus a CT of <40 for ptxS1, the gene that codes for pertussis toxin. It should be noted that IS481 is a very sensitive probe as *B. pertussis* carries multiple gene copies per organism. By contrast, there is usually only one copy of ptxS1 per bacillus, making it highly specific but insensitive.^50,51^ We then expanded testing for IS481 and ptxS1 to all of the other samples within those eight mother/infant pairs.

#### Systematic analysis of the full data set

We focus here on IS481 qPCR CT values. We did not include ptxS1 in these analyses due to its lack of sensitivity. While this introduces the possibility that some IS481 detections were due to a species other than *B. pertussis*, we expect a minimal impact of rare *B. pertussis* false positives on our findings. As the duration of study participation is somewhat variable between subjects (**Figure S1-S3**), we focus on subjects with at least 4 NP samples.

##### Temporal Analysis

We first conduct an exploratory data analysis of IS481 CT values over time. We group assays into arbitrary ranges of CT values such that each range of decreasing CT value (greater target) contains fewer samples than the previous interval. We then use a set of generalized additive models (GAMs) to describe the relative frequency of samples in each CT range over the course of the study (binomial link function, smoothed by calendar date using cubic regression splines with shrinkage, one model per stratum). We use these models to visually compare the relative frequency and timing of IS481 signal intensity.

##### Evidence for Infection

For each participant, we compute a summary measure of their IS481 CT values across the course of the study, which we refer to as the *evidence for infection* (EFI). To compute the EFI, we first compute the reverse cumulative distributions (RCD) of CT values over all samples in the study (for mothers and infants separately). Emphasizing that the CT is roughly equivalent to the inverse of pathogen density, from these RCDs, each IS481 assay is now associated with a probability describing its rarity in the study, ranging from 1 (not detected after 45 cycles) to 0 (lowest CT value in the study). For each subject, we then compute the geometric mean RCD probability of that subject’s assays. One minus this mean proportion yields the *evidence for infection* (EFI) in this subject during the study period. Conceptually, zero detecting assays (EFI=0, all N.D.) indicates no evidence for infection, whereas an EFI approaching one indicates strong evidence for infection at some point during the study (but does not provide information about the timing of infection).

We also assess the number of detecting assays per subject. We consider individuals with 3 or more detecting assays as very likely to have experienced *Bordetella* infection during the study. We then identify the minimum EFI of these individuals, and take this value as a threshold delineating *strong* evidence for infection (EFI ≥ threshold). Note that, since our false detection rate is bounded above by approximately 10% (assuming all detections were false, **Table S1**), our false classification rate given 3 or more detecting assays must be less than 10%^3^ = 0.1%. Subjects with EFI below this threshold but greater than zero we consider as having *weak* evidence for infection, while subjects with EFI equal to zero have no evidence for infection (all N.D.). Thus, each mother and infant in the analysis set is assigned into one of three mutually exclusive categories: no evidence, weak evidence, and strong evidence. As with EFI, these categories contain no information about the timing of infection.

We next assess the correspondence of EFI within mother/infant pairs. We compute the frequency of EFI category (none, weak, or strong for mothers versus infants), along with Pearson residuals from a model assuming independent association between mothers and infants. The residuals effectively rate the strength of association between the mother/infant pairs. We also test the dependence of our results on the specific threshold used to determine EFI strength. To do this, we rerun the above analysis using all individuals with either 2 or 4 detecting assays as sensitivity analyses to explore the impact of varying the EFI threshold (**Figure S5)**.

We also assess the correspondence of EFI with clinical symptoms of pertussis and antibiotic use. Here we categorize symptoms as mild (cough or coryza) or moderate to severe (all other symptoms indicated by the modified Preziosi Scale^52^). We also categorize individuals according to the worst symptoms observed at any point during the study.^53^ Likewise, participants are categorized according to whether antibiotics were used at any point during the study. This correspondence analysis was conducted separately for infants and mothers.

## Supporting information

STROBE checklist

## Data Availability

Data will be provided in a publicly available repository upon publication

## ACKNOWLEDGMENTS AND FUNDING SOURCES

We wish to thank the Lusaka lab team who generated the results for this analysis: Caitriona Murphy; Ruth Nkazwe; Chilufya Chikoti; and Baron Yankonde.

The lab testing and subsequent analyses for this paper were supported by a grant from the National Institutes of Health/National Institute of Allergies and Infectious Diseases (R01AI133080). Funding for the initial SAMIPS study that allowed us to create the sample library itself was through the generous support of the Bill & Melinda Gates Foundation (OPP1105094).

