## Supplemental Figures and Tables for "Asymptomatic *Bordetella pertussis* infections in young African infants and their mothers identified within a longitudinal cohort"

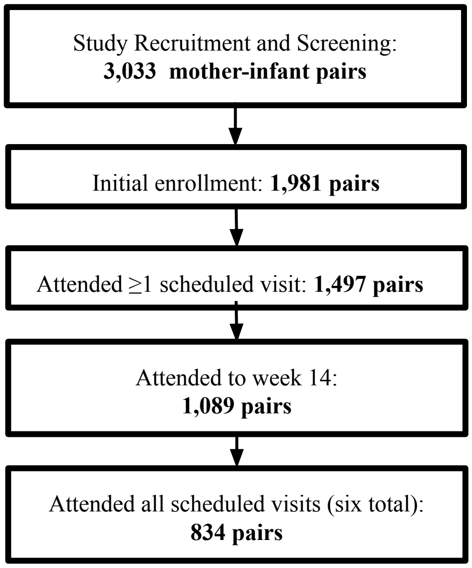


Figure S1: Study profile of birth cohort enrollment and attendance. Beyond eligibility and initial screening, the sole cause of cohort attrition was failure to attend one or more scheduled clinic visits. For eligibility and enrollment details, please see Gill et al. Incidence of Severe and Nonsevere Pertussis Among HIV-Exposed and -Unexposed Zambian Infants, CID 2016.

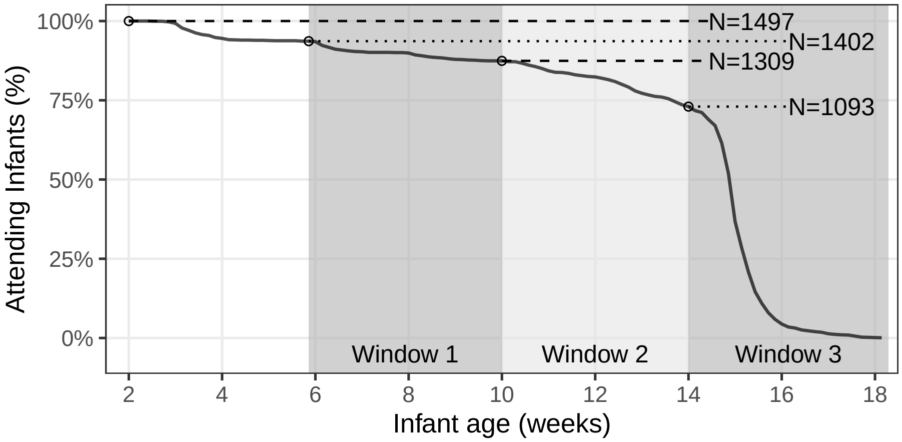


Figure S2: Percent of attending infants by age of last attendance. Shaded regions show target age windows of DTP doses 1-3. Horizontal lines and text shows number of infants attending up to marked ages (excluding non-attending infants): beginning of study enrollment, and 41, 70, and 98 days (earliest timely administration of DTP doses 1-3, respectively). Study profile: initial enrollment, 1981 mother/infant pairs; attended $\geq$ 1 scheduled visit, 1497 pairs; attended scheduled visit in window 3 (i.e., at study conclusion), 1093 pairs; attended all six scheduled visits, 834 pairs.

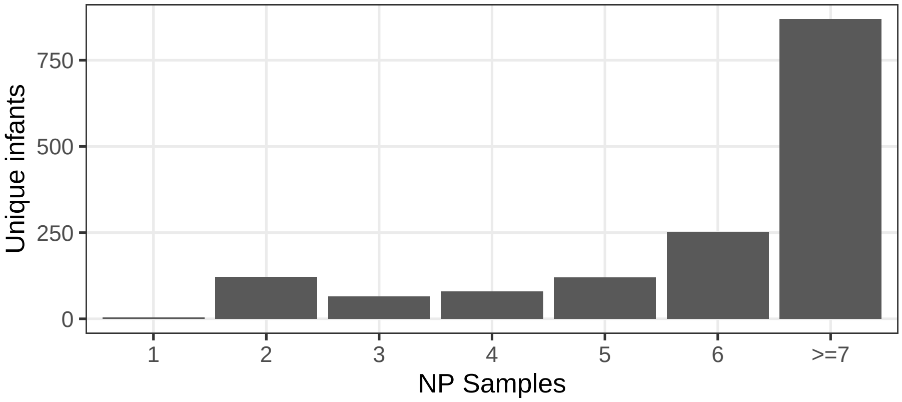


Figure S3: NP samples per infant, showing number of attending infants with each sample count (including enrollment and unscheduled visits). A majority of infants (834/1,497) attended all scheduled visits.

Table S1: Summary of pertussis assays (qPCR, IS481) for infants and mothers, showing the frequency and percent of nasopharyngeal (NP) samples in each cycle threshold (Ct) interval.

| Ct Interval | Mother | Infant |
|---|---|---|
| [16,35) | 10 (0.11%) | 16 (0.18%) |
| [35,41) | 96 (1.1%) | 114 (1.3%) |
| [41,43) | 230 (2.6%) | 223 (2.6%) |
| [43,45] | 482 (5.5%) | 390 (4.5%) |
| N.D. | 7901 ( 91%) | 7980 ( 91%) |

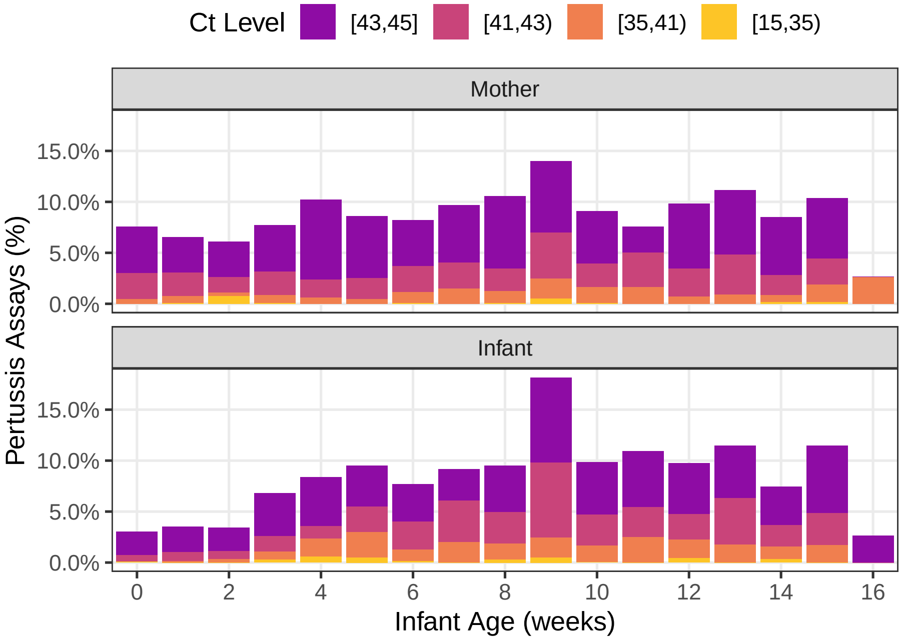


Figure S4: Percent of IS481 assays in each Ct Interval by infant age (aggregated by week). Weeks with < 20 assays are omitted for clarity.

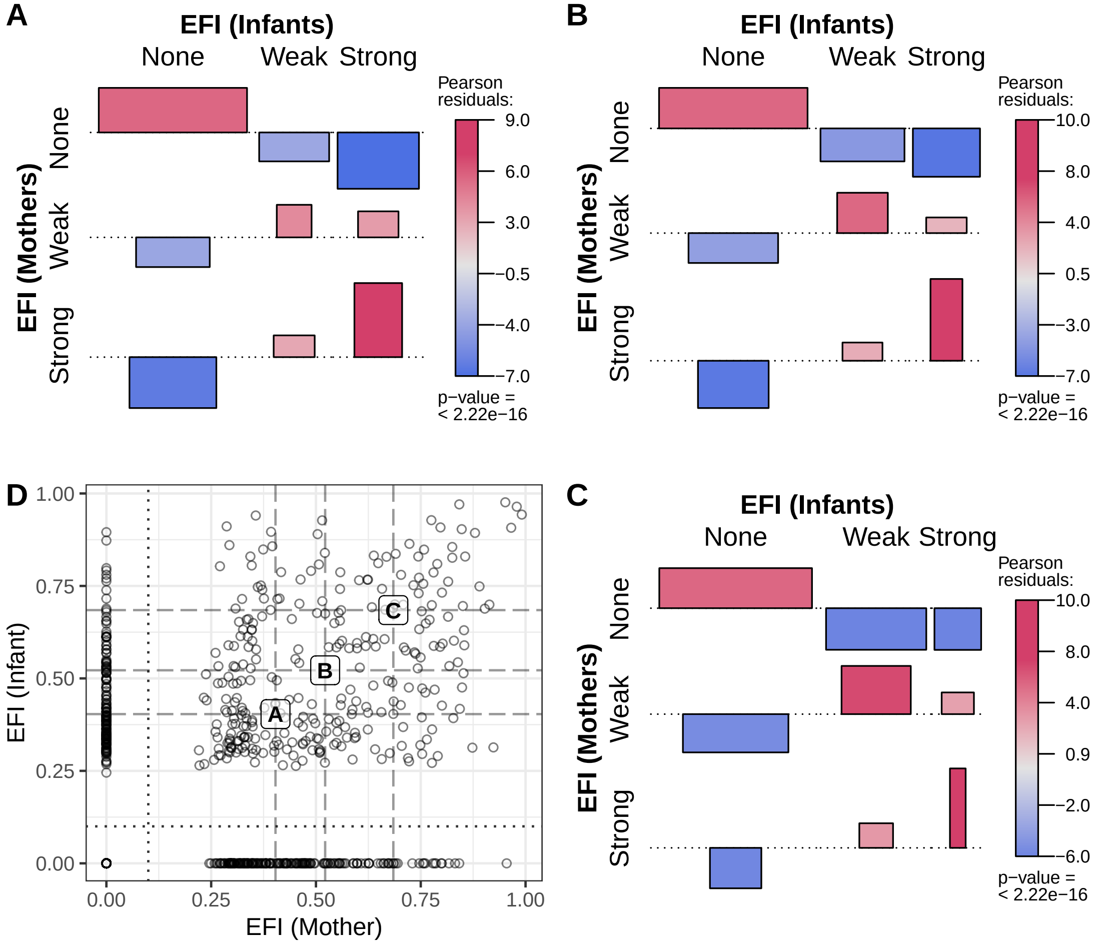


Figure S5: As in Figure 3, varying the threshold of `Strong` EFI (dashed line) to include all individuals with ≥2, 3, or 4 detecting assays (A-C, respectively). As the threshold increases from A to C, strong EFI is observed in fewer individuals (and weak EFI in more), but the pattern of association between mothers and infants remains largely unchanged.

Table S2: Details for subjects shown in Figure 1, including the number of NP samples, number of samples with detected IS481, and EFI, along with summary of pertussis symptoms and antibiotics use.

| Subject | ID | Samples | Detected | EFI | Symptoms | Antibiotics |
|---|---|---|---|---|---|---|
| A Infant | 126 | 11 | 3 (27.3%) | Strong (0.68) | Mod/Sev | Yes |
| A Mother | 126 | 11 | 3 (27.3%) | Strong (0.52) | Mod/Sev | Yes |
| B Infant | 269 | 7 | 4 (57.1%) | Strong (0.89) | Minimal | Yes |
| B Mother | 269 | 8 | 1 (12.5%) | Weak (0.50) | None | No |
| C Infant | 434 | 7 | 3 (42.9%) | Strong (0.84) | Minimal | No |
| C Mother | 434 | 7 | 2 (28.6%) | Weak (0.52) | None | No |
| D Infant | 474 | 9 | 4 (44.4%) | Strong (0.94) | Mod/Sev | Yes |
| D Mother | 474 | 9 | 7 (77.8%) | Strong (0.99) | Mod/Sev | Yes |
| E Infant | 573 | 7 | 4 (57.1%) | Strong (0.87) | Minimal | Yes |
| E Mother | 573 | 7 | 0 (0.0%) | None (0.00) | Minimal | No |
| F Infant | 579 | 7 | 4 (57.1%) | Strong (0.90) | Minimal | No |
| F Mother | 579 | 7 | 3 (42.9%) | Strong (0.85) | Minimal | No |
| G Infant | 691 | 7 | 4 (57.1%) | Strong (0.96) | Mod/Sev | Yes |
| G Mother | 691 | 7 | 6 (85.7%) | Strong (0.98) | None | No |
| H Infant | 752 | 9 | 7 (77.8%) | Strong (0.98) | Mod/Sev | Yes |
| H Mother | 752 | 9 | 7 (77.8%) | Strong (0.95) | None | No |
